# Artificial Intelligence for the Diagnosis and Treatment of Diabetes Kidney Disease: a systematic review

**DOI:** 10.1101/2021.10.10.21264813

**Authors:** Shams Mohammad Abrar

## Abstract

**Background:** Diabetic nephropathy (DN) is a serious microvascular complication that affects 40% of diabetes patients. In the last decade, artificial intelligence (AI) has been widely used in both structured and unstructured clinical data to improve the treatment of patients/potential patients with DN.

**Methods:** This systematic review aims to cover all applications of AI in the clinical use of DN or related topics. Studies were searched in four open-access databases (Pubmed, IEEE Xplore, DBLP Computer Science Bibliography, and ACM digital library). Finally, the author manually searched the reference list of included studies in the study for additional relevant articles.

**Results:** Finally, a total of 24 original peers reviewed articles were included in this study. Through a manual data extraction, the summary of key information such as applied AI algorithm, main outcomes, performance evaluation etc. was taken. Then the included studies underwent a quality assessment criterion, assessing the reproducibility, generalizability etc. Most of the included studies revealed that the AI frameworks outperformed conventional statistical methods. A summary of the limitations, such as lack of data availability or external validation of the framework, in the included studies, was also included.

**Conclusion:** The rapid advancement of the AI framework and the exponential data generation in healthcare can be utilized and applied in clinical practices. The aid of AI can be instrumental in the treatment of DN.

## 1. Introduction

Diabetes Mellitus (DM), a devastating incurable health complication, is caused due to the incapability of beta cells (β cells) in the pancreas to produce adequate effective insulin for the body to utilize - leading to chronic high levels of blood glucose (1). Globally diabetes is considered a major public threat. According to the International Diabetes Federation (IDF), every eleventh person on the planet (463 million, 95% confidence interval: 361-601 million) has diabetes, and half of these patients are undiagnosed due to the disease’s complex pathogenesis. Furthermore, diabetes accounts for ten per cent of global health expenditure (USD 760 billion) (2). Several factors have been linked to the onset of DM in patients. Inactive lifestyles, obesity, a lack of awareness, and other factors have all been linked to the recent global increase in diabetes (3).

DM, on the other hand, results in a variety of macro and microvascular complications as a result of hyperglycemia or hypoglycemia due to the lack of blood glucose (BG) control. Diabetic Nephropathy (DN)/ diabetic kidney disease (DKD) is one of the microvascular complications, which is the most common cause of end-stage renal disease in the world (4-7). DN is caused due to increased urine albumin excretion and/or impairment. It is characterized by gradual loss of kidney function from hyperfiltration at the early stage (8). The principal risk factors involving DKD are hyperglycemia, hypertension and genetic predisposition. To diagnose DN early on, it is recommended to screen microalbuminuria five years after diagnosis for type 1 diabetes (T1D) and yearly screening for type 2 diabetes (T2D) (9). However, the amount of DM patients being affected by kidney disease is rising rapidly. Prevalence of microalbuminuria is seen in about one-third of DM patients after fifteen years of disease duration, and almost half of them develop DN. Often DN occurs in patients of a certain age group or demographics. It is found that DKD is mostly common among elderly patients, very unlikely to develop among patients with less than one decade of diabetes duration (10). Meanwhile, the risk of developing DKD is higher among patients from developing countries compared to developed countries (26). The cost of bearing dialysis and other casualties is very high. As DN is more prevalent in developing countries, it almost becomes impossible for the patient’s family to bear the expenses. Meanwhile, the mortality rate of DN patients is very high, and early diagnosis or adequate treatment is necessary. Around 10% of the death in T2D is due to kidney failure (11).

Despite growing concerns and preventative measures of DKD, the rate of chronic kidney disease (CKD) among DM patients remains unchanged as of 20 years ago. Even though DN might be controlled by the maintenance of blood glucose, blood pressure, blood cholesterol and widespread use of renin-angiotensin-aldosterone system inhibitors (9), it only works at the early stages. On the other hand, the progression of DKD of individual patients shows a large variation due to the complex heterogeneous nature of the disease (41). At later stages of DN, when CKD is well developed, it is harder to control (12). It is found that early diagnosis of microalbuminuria among DM patients and monitoring will postpone or prevent overt nephropathy (13,14). As a result of risk verification or early diagnosis of DN a large sum of the socioeconomic burden suppressed on the patients and their families can be avoided - where the further progression of DN could be stopped (12).

With the help of machine learning (ML), a branch of artificial intelligence (AI) diagnosis, prognosis and clinical management of DKD patients without direct human intervention is possible. These algorithms are capable of extracting clinically relevant information from medical data with great accuracy (8). Without being explicitly programmed, machine learning algorithms can produce non-linear correlations and patterns from raw data (15). Deep learning (DL), a branch of machine learning that is aided by increased computational capabilities, entails training a large number of neural networks with many layers on large data sets. As a result, deep learning algorithms are useful for image classification in medical datasets (11). This systematic review covers the application of AI in DKD, which includes the early detection of DN to the management of DN through the application of AI.

Several systematic reviews have examined the use of AI in diabetes. Zhu et al. investigated the use of DL in DM, searching multiple databases and obtaining 40 studies (16). A total of 107 peer-reviewed research articles were included in a study by Jyotismita Chaki et al. on the use of AI in the detection and self-management of diabetic patients (17). Ioannis Kavakoitis et al. discussed the use of machine learning and data mining in diabetes care (18). These studies, on the other hand, cover the broader application of AI in DM patients. They are not focused on any specific complications. Meanwhile, Katrine et al. conducted a systematic review on the use of DL algorithms in the screening for diabetic retinopathy (DR), evaluating the diagnostic performance of DL algorithms. Diabetic retinopathy is the most common diabetic microvascular complication. The study included a total of 20 articles (19). Miguel Tejedor et al. discussed the use of reinforcement learning, a type of machine learning in which the agent learns in an interactive environment through trial and error and feedback, in diabetic glucose control (20). A systematic review with meta-analysis was performed by Md Mohaimenul et al. about the detection of DR from retinal fundus photographs, where a total of 20 studies were included (21). However, there has yet to be a systematic review of AI’s application in DN. This research aims to fill that void. This systematic review includes studies that use AI algorithms to address the early detection of DN, the prognosis of DN, DN management, and other related topics.

## 2. Overview of Big Data analytics and AI

Researchers have been craving, since the mid-twentieth century, for a support system that can make clinical decisions while processing growing quantities of clinical data (22). As the medical records are often high dimensional, heterogeneous and sparse they are often not utilized properly in clinical practices (9). Applied biology is advancing to the era of ‘big data’ due to the remarkable advances in biotechnology. The principal reason behind the establishment of big data in medicine is high throughput sequencing due to the advancement of biotechnology (23,24). Myriads of large-scale and real-world data are generated from the likes of hospital records, patients’ medical records, biomedical research etc. However, to utilize the data generated requires proper management and analysis (25). The proper application of big data in healthcare is executed by extracting patterns and trends thus creating models from the data. Consequently, big data analytics will yield more throughput insightful diagnoses, prognosis and most clinically effective treatments in a cheaper method, and it will improve healthcare marginally (26). An abundance of clinical data and information regarding diabetes patients is already available and it is increasing at an exponential rate (27).

Artificial Intelligence (AI) is a branch of computer science that aims to mimic the cognitive function of human beings. There are two main branches of AI: virtual and physical. The physical branch covers the aspects of robotics, which can assist in surgery in clinical practice. On the other hand, the virtual branch of AI includes the likes of ML. This informatics branch of AI is capable of assisting physicians in clinical diagnosis, treatment decisions etc. (28). As a result of the ‘big data’ era’s emergence, AI brings forth permanent change in the healthcare system. In addition, AI applies to both structured and unstructured data. ML and Natural Language Processing (NLP) is applied in structured and unstructured data respectively (29). The raw data available from EHR usually consisted of structured data. Thus, ML algorithms are mostly used as predictive performance models in different studies. ML is classified into three different categories: supervised learning, unsupervised learning and reinforcement learning. In both supervised and unsupervised learning, the system infers a function from labelled and unlabeled data respectively. ML workflow often involves data harmonization, preparation learning, model fitting and evaluation (30). Figure 1 demonstrates the learning process of both supervised and unsupervised learning (31). The theoretical construct of ML models gives them an edge over conventional statistical models. As they are efficient at analyzing large complex datasets, while identifying hidden associations ML is proved to be superior. Thus, over the recent decade, ML has evolved dramatically, and it is commonly used in clinical practices (16).

**Figure 1:**
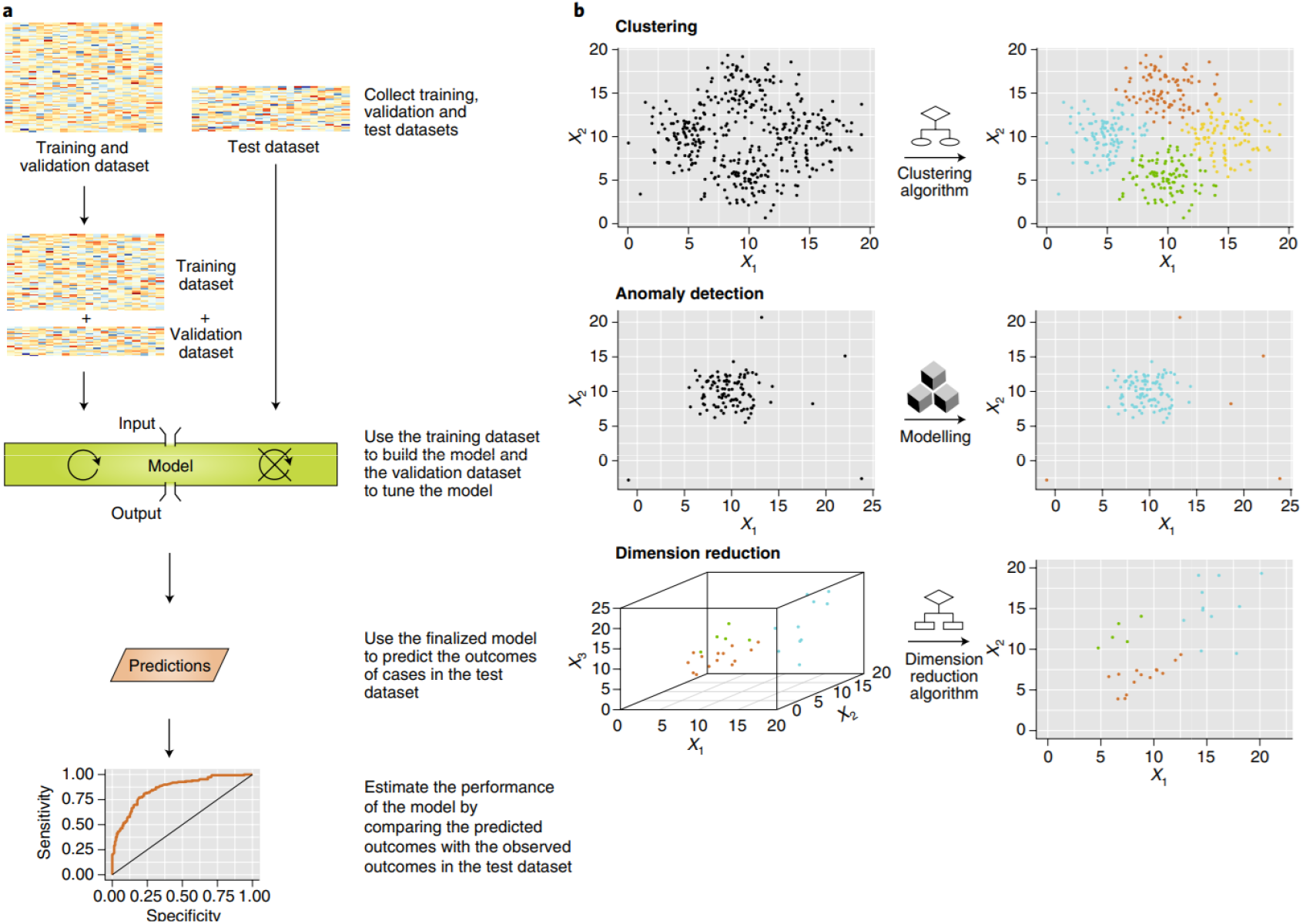
Supervised and unsupervised machine learning. **a**, General workflow of supervised machine-learning approaches. First, training and test datasets are collected. Next, part of the training set is used to build the prediction model, and the other part to tune and validate the model (circular arrow). After the machine-learning model is finalized (crossed-out circular arrow), the established model is used to generate predictions on the test dataset and the model’s performance is estimated by comparing the predicted outcomes with the observed outcomes for the test dataset. **b**, Unsupervised machine learning includes clustering, anomaly detection and dimensionality reduction. Clustering algorithms group data points with similar measurements into clusters. Anomaly detection identifies outliers in the dataset. Dimensionality reduction reduces the number of random variables used to describe the data; for example, by representing an image with thousands of parameters as a smaller vector of summary features. The resulting summary vector preserves the important information in the raw data; for example, summary vectors from similar images will bear more resemblance than those obtained from irrelevant images Note: Descriptive phrase that serves as title and description. Reprinted from “Artificial Intelligence in Healthcare” by Ognjanovic I, Year 2020, Stud Health Technol Inform, *Volume*(issue), doi: 10.3233/SHTI200677. PMID: 32990674 (7).

On the other hand, DL is a subfield of ML. The field of DL has advanced massively in the ability of machines to understand and data including the likes of images, language and speech. In contrast to ML, DL is a form of representation learning composed of numerous layers - gaining the ability to learn highly complex functions (33). DL algorithms such as Convolutional Neural Network (CNN) is designed to process data that exhibits natural spatial invariances. Therefore, DL models have achieved the accuracy of physicians in identifying cardiovascular risk from fundus images (34). Compared to the traditional Artificial Neural Networks (ANN), which are limited to three layers and trained to obtain supervised representation for specific tasks only (35), DL layers of features are not designed by anyone, but they are learned from data using a general-purpose learning procedure. The inputs of deep neural networks are processed in a non-linear manner to activate the nodes in hidden layers to learn the ‘deep structures and representations. Next, the final representation is utilized in a supervised layer to fine-tune the network using backpropagation algorithms toward representations, which are optimized for an end-to-end task (36).

## 2. Methods

A qualitative systematic review of the application of AI for the diagnosis, prognosis and management of DN was conducted. This systematic review was conducted by following the reporting checklist of Preferred Reporting Items for Systematic Reviews and Meta-Analysis (PRISMA 2020) (37). Consequently, comprehensive literature was undertaken with extensive efforts to identify the articles. This review was conducted from 2020 to 2021 (considering all articles published from 1970 - 2021), and the databases searched include PubMed, DBLP Computer Science Bibliography, IEEE Xplore and ACM digital library. The four databases used are interfaces that do not require institutional subscriptions unlike other databases such as Ovid, Scopus and Web of Science. The purpose of using open-access databases is to enhance the reproducibility of the search results (16). The last search performed was on 12th July 2021.

### 2.1 Search Strategies

The steps of searches were performed using related keywords combinations based on mesh terms. For this systematic review, related keywords of diabetes and nephropathy and AI were combined using Boolean operators (AND/OR). The search strategy is given in Table-1.

**Table-1:** The databases and the search strategy employed

|  |  |
| --- | --- |
|  | Search Strategies |
| Database | PubMed, ACM Digital Library, IEEE Xplore, DBLP Computer Science Bibliography |
| #1 | diabet* OR DM OR T1D OR T2D OR Hyperglycemia OR “glucose intolerance” |
| #2 | Nephropathy OR “kidney disease” OR kidney OR Renal |
| #3 | “Artificial Intelligence” OR “machine learning” OR “deep learning” |
| Search | #1 AND #2 AND #3 |

At the outset, the duplicate articles from different sources were excluded from the initial collection of the obtained articles. The records were screened a total of three times by the author, to avoid any biases in the systematic review. Additional potentially eligible articles were searched manually by the author by screening the reference list of included articles.

### 2.2 Inclusion and Exclusion Criteria

The most relevant and potential articles from the initial screening were kept for full-text review. To be included in the review, the articles had to fulfil certain criteria. The study must be in English and peer-reviewed. In addition, application of AI algorithms with performance evaluation is necessary. On the other hand, the study must be applicable for Diabetic patients with potential kidney disease or persisting kidney disease. Any study excluding the application of AI in diabetes and kidney disease were not included. If the AI algorithm only applies to DM but is not related to nephropathy is excluded - vice versa is also applicable. In addition, other types of study such as letters to editors, posters and short communications were excluded. As well, the included articles had described thoroughly the algorithms used and the data processing of datasets. Traditional methods used only in DN are excluded. The author, himself, had thoroughly examined the selected articles by screening the article titles and abstracts to identify the eligible articles. If the article is not relevant to the topic at all then it is excluded at the initial phase which is conducted by screening the title and abstract.

### 2.3 Data extraction

From the final collection of the studies, key information was extracted to assess the application of AI algorithms. The data extraction was divided into 10 different categories. The categories and their information are given below:

1. Study information: the first author’s name, year, country.
2. Cases: summary of the specific application in different cases i.e., detection of DN, management of DN etc. If applicable the type of diabetes of the cohort was mentioned as in T1D or T2D.
3. Data sources: The name and description of the dataset’s source is obtained and mentioned. It can be an EHR dataset from a regional hospital to national surveys.
4. Research type: The type of study conducted, whether it was an observational study or experimental study, is mentioned in this category.
5. Baseline characteristics: The clinical variables used to form the ML models were mentioned here. For DL algorithms the type of image taken along with basic characteristics is mentioned.
6. Cohort size/sample size: The population included in the cohort studies is mentioned. On the other hand, the sample size for other studies is mentioned here.
7. AI algorithm: In this category, all of the ML or DL algorithms used are mentioned.
8. Development process: The size of the training and testing dataset is mentioned here.
9. Performance assessment: The evaluation metrics of AI algorithms are included here. Some of the performance assessment techniques include accuracy test, sensitivity, specificity, Cohen’s k, the area under the curve (AUC) etc.
10. Limitation: The limitations of the included studies, mostly mentioned by the authors themselves, are included in this category.

### 2.4. Quality Assessment

Studies were assessed from the following categories: limits in current non-machine learning tools, feature engineer methods employed in the AI algorithm before training the dataset, the hyperparameters used in the study, valid methods to overcome overfitting and the external validation of the data. The quality assessment table of the studies consisted of binary answers of yes or no.

## 3. Results

After obtaining the results from the four databases, the duplicate articles were excluded. Then a manual inspection by the author was conducted to evaluate the remaining articles based on inclusion criteria. From the four databases a total of 694 (IEEE = 91, PubMed = 202, ACM Digital Library = 393, DBLP = 8) studies were obtained, and 11 of them were marked as ineligible by endnote. In addition, a total of 24 duplicate studies were found. The remaining 659 studies were manually screened by the author, and they were evaluated based on the inclusion and exclusion criteria. Consequently, a total of 146 studies were retrieved for eligibility assessment via full-text inspection - all of these studies either included diabetes patients and/or kidney patients and/or AI algorithms. In the end, a total of 23 articles met the inclusion criteria. The references cited in those 23 studies were screened by the author himself. Among the 831 references listed only one of the studies met the complete inclusion criteria. The details of the selected articles are presented in Table-3.

**Table-2:**
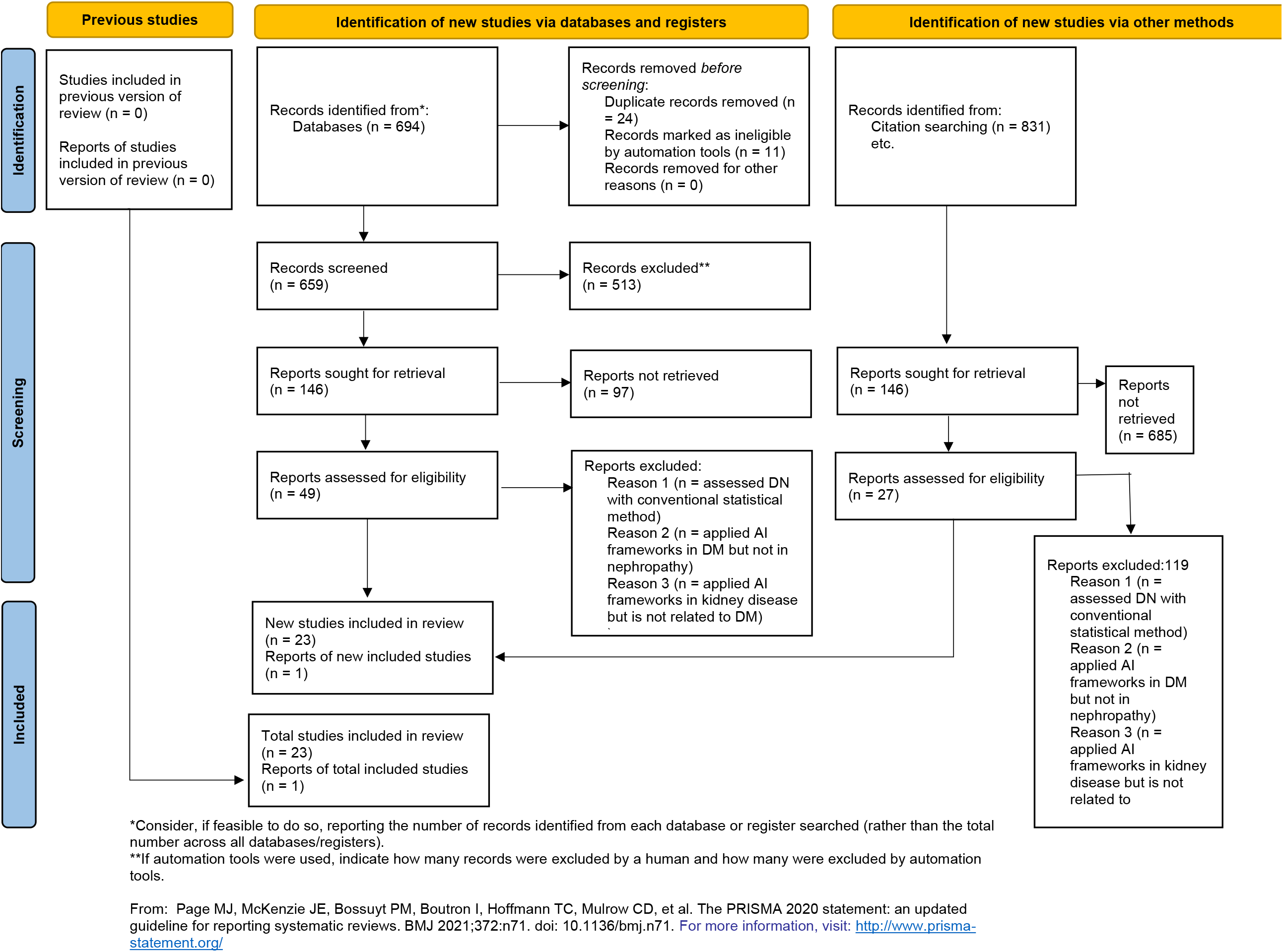
PRISMA flow diagram for updated systematic reviews.

**Table-3:**
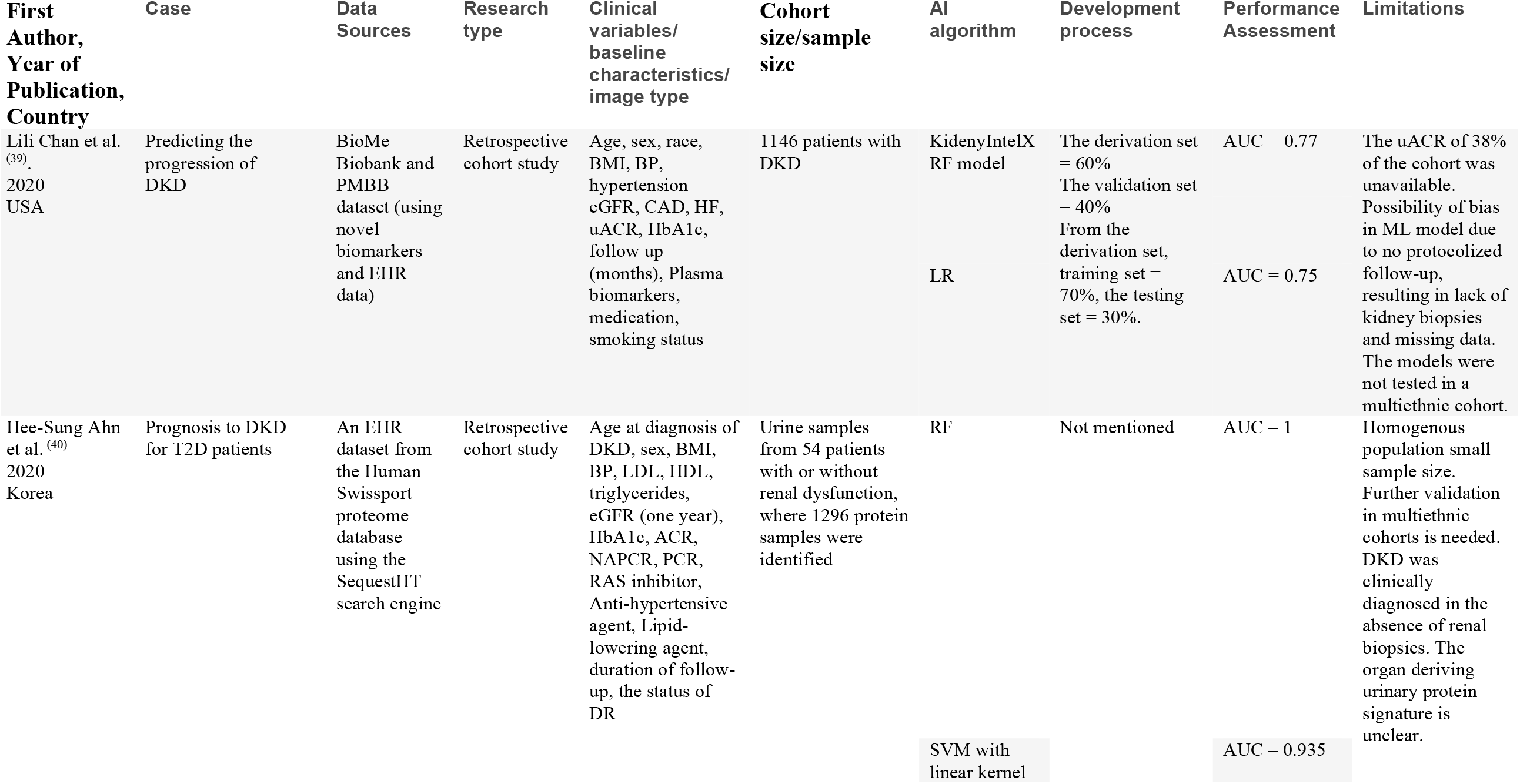
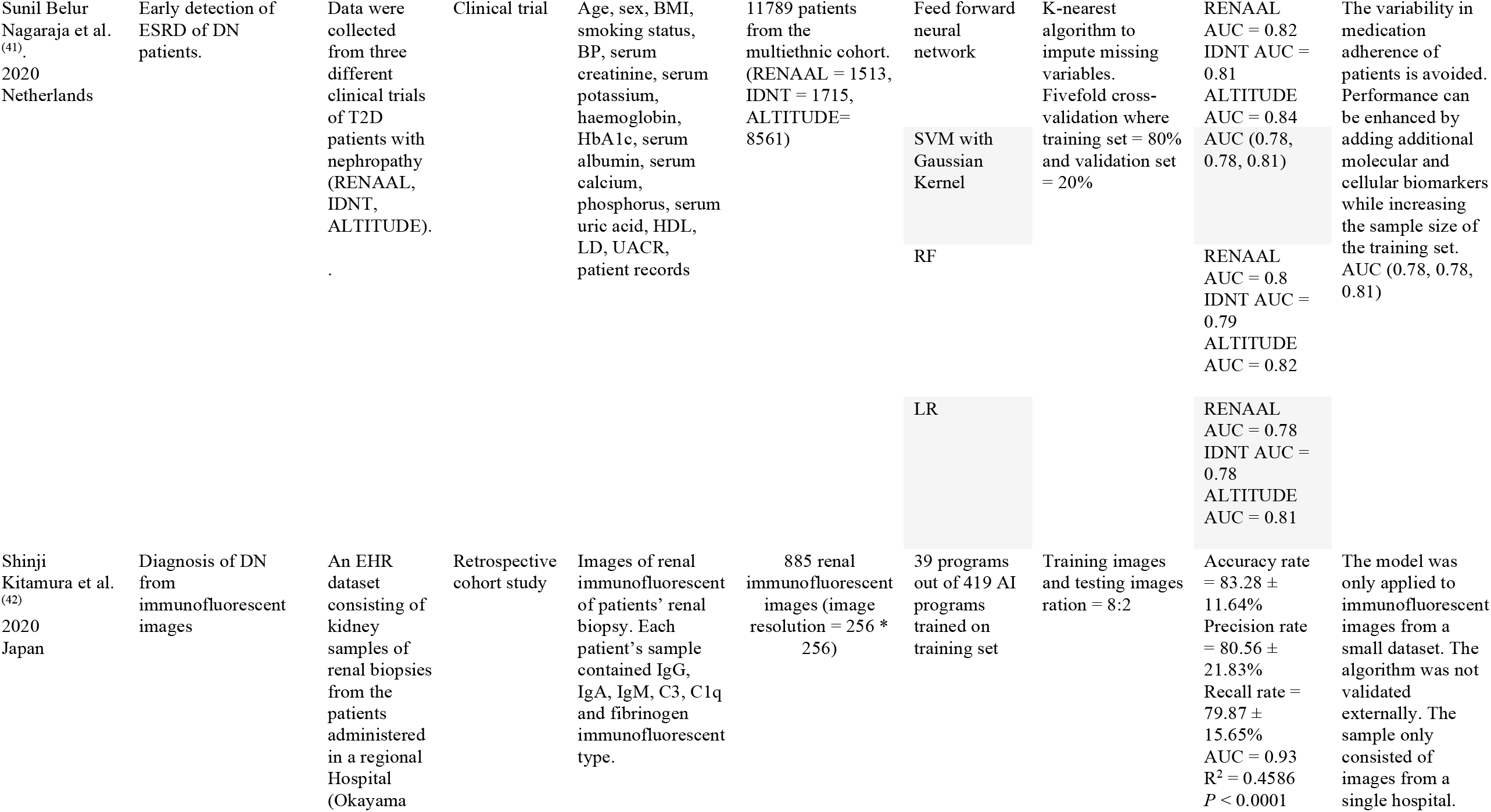
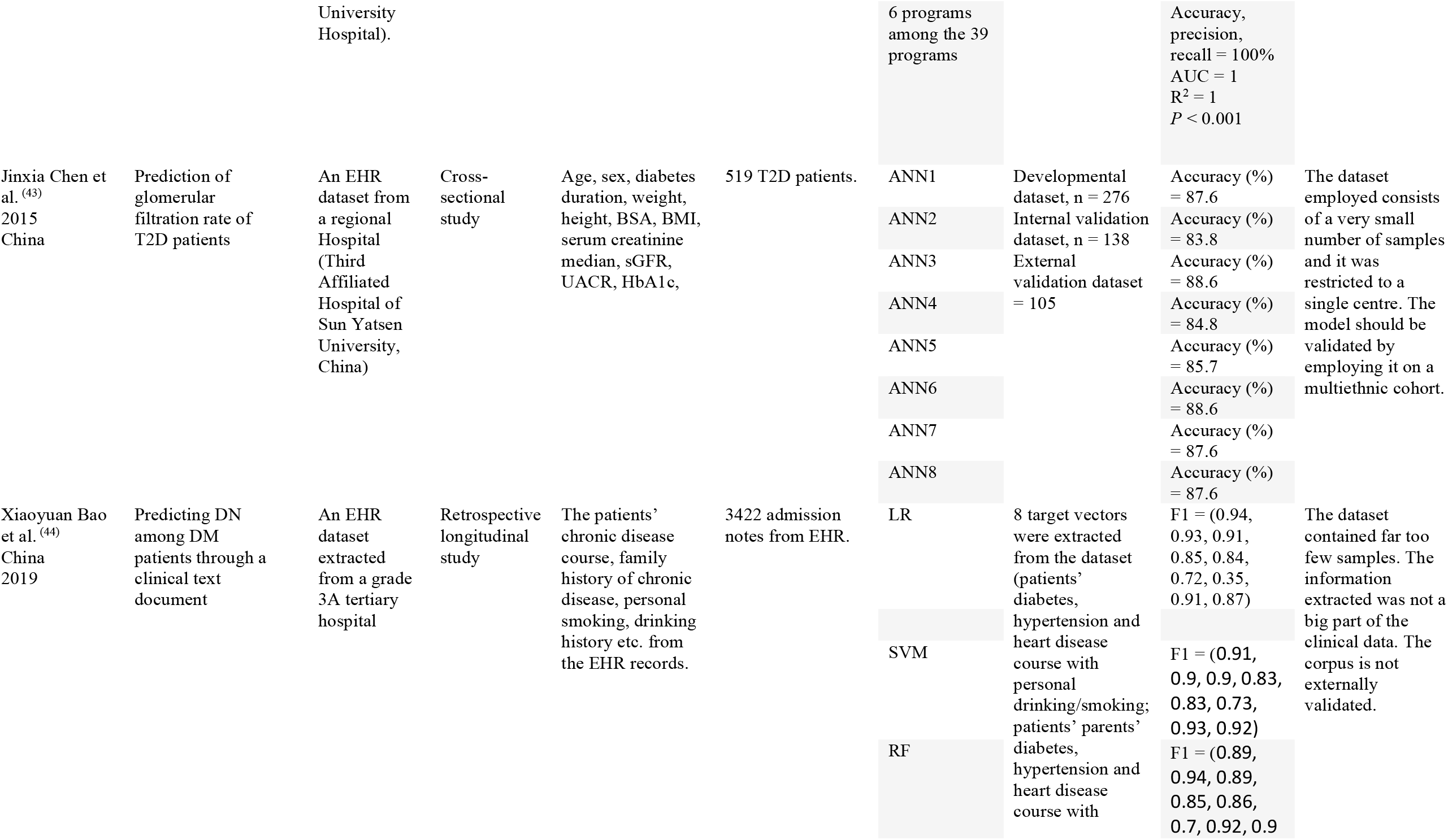
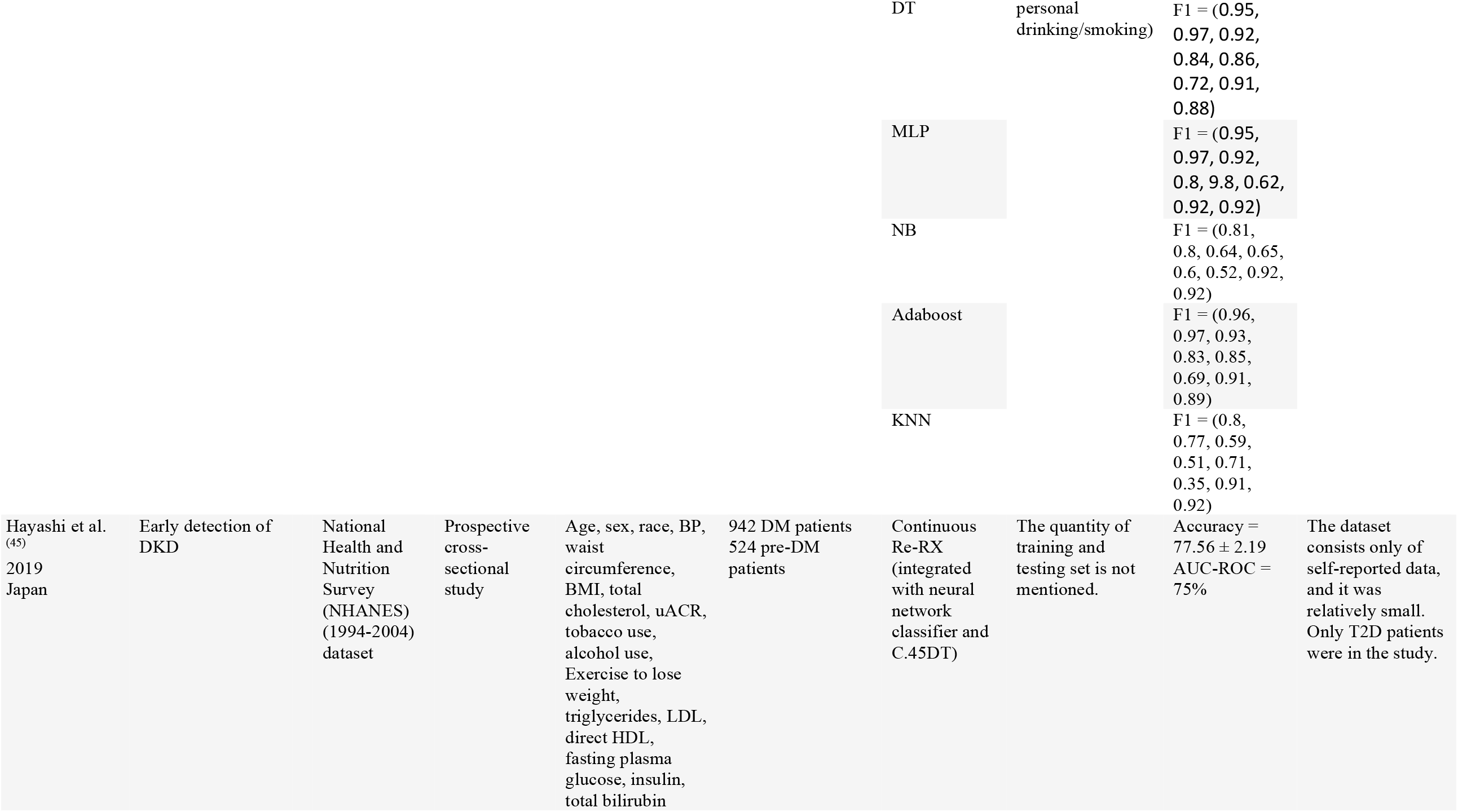
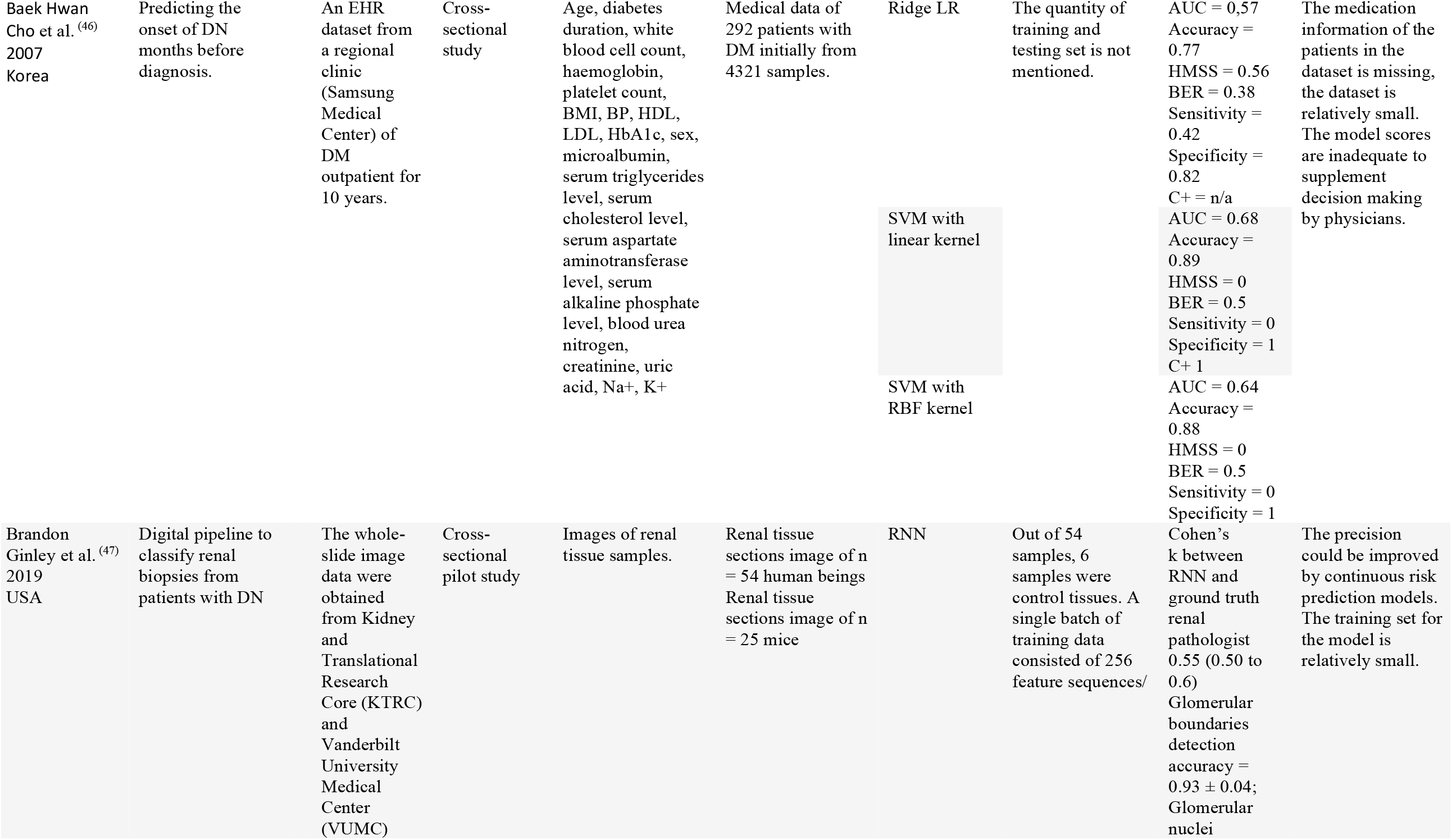
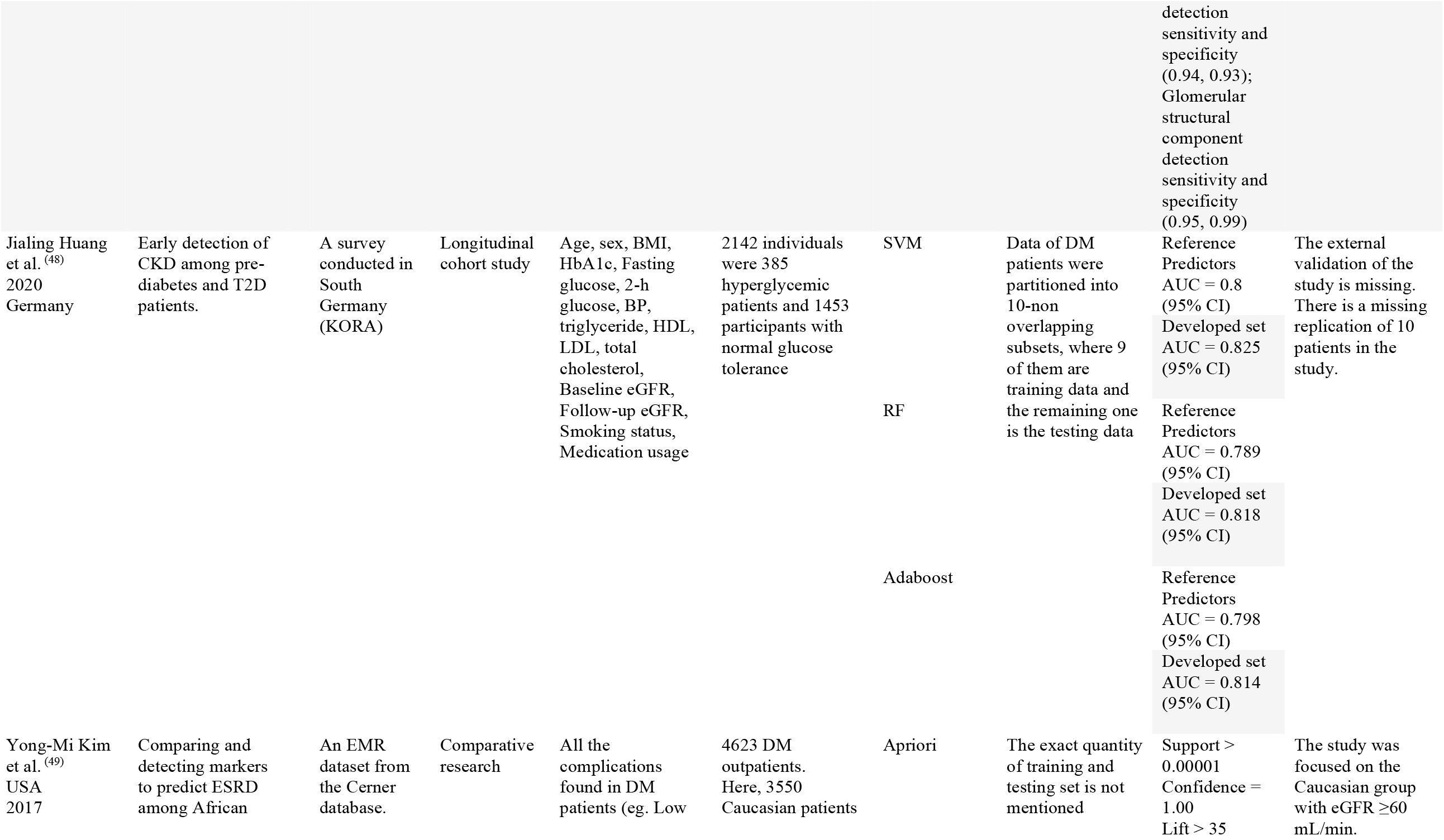
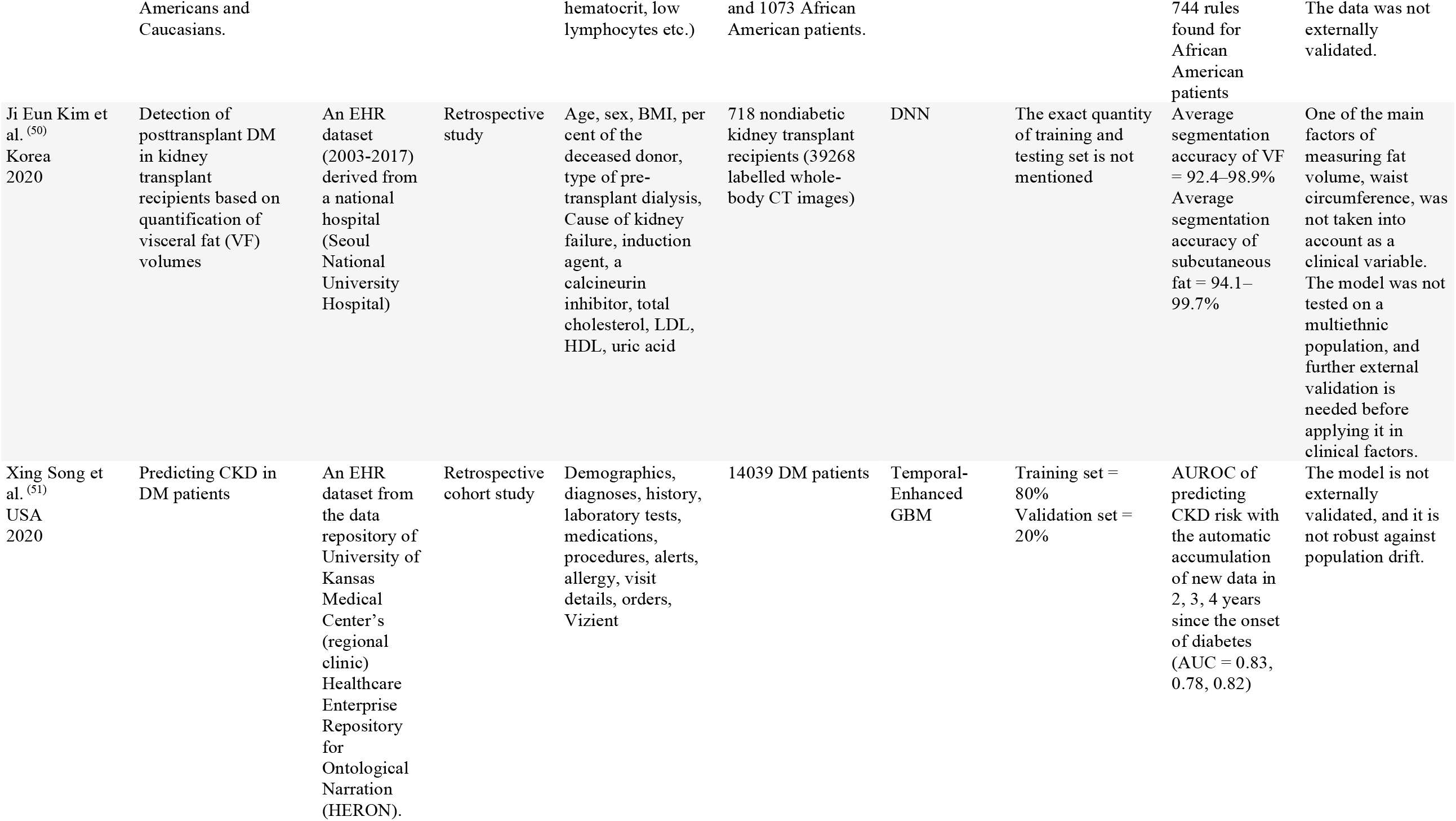
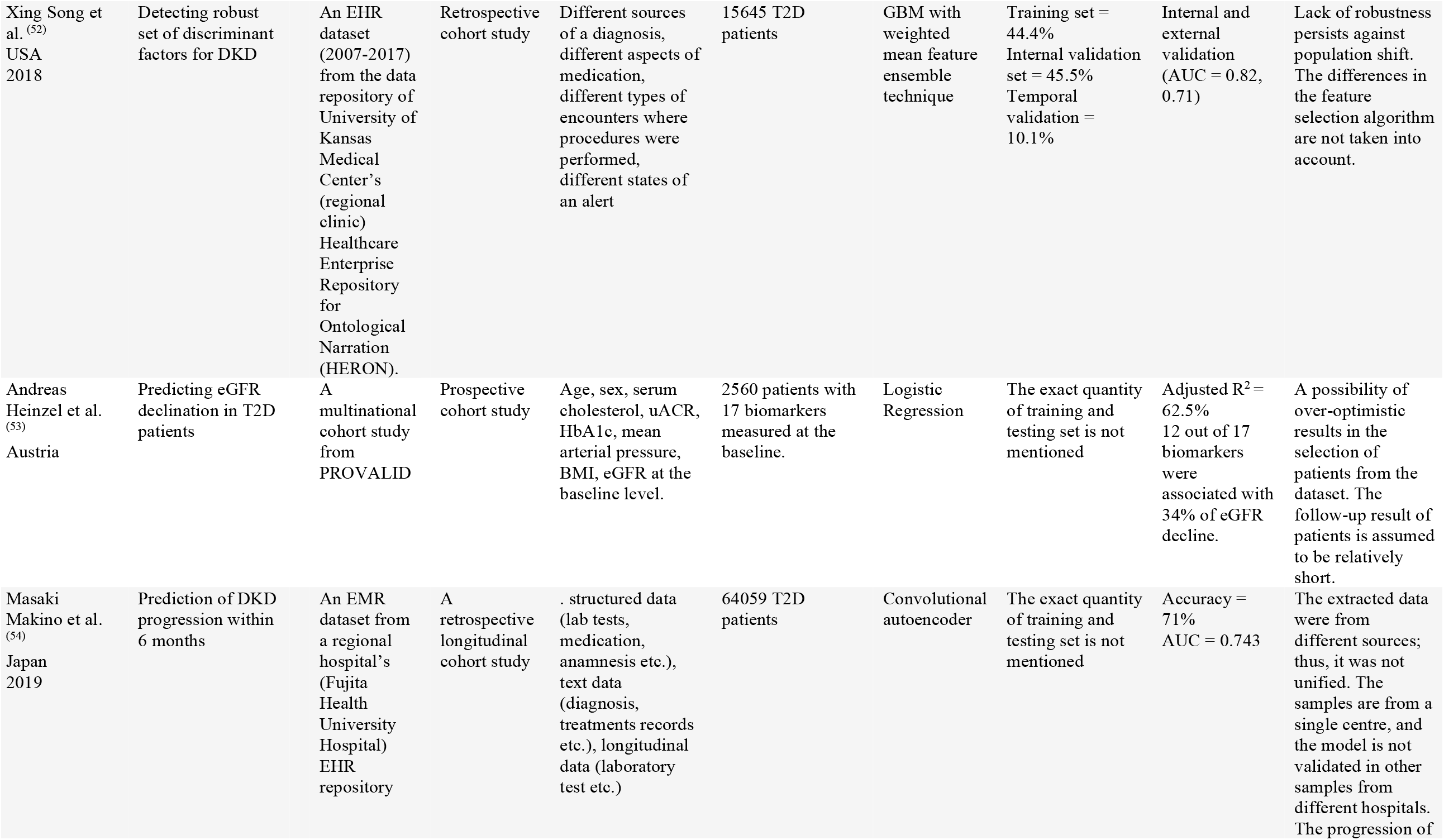
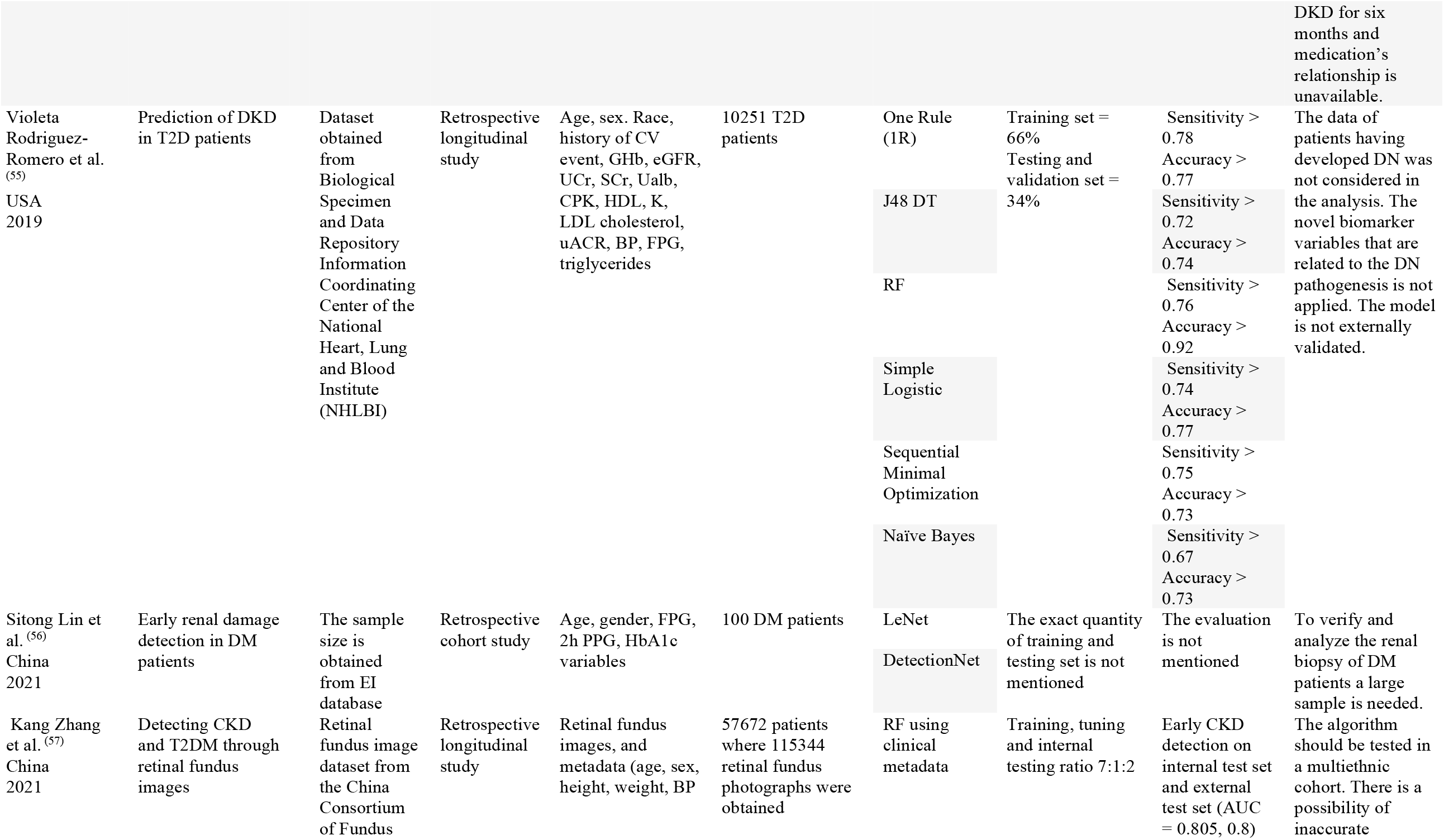
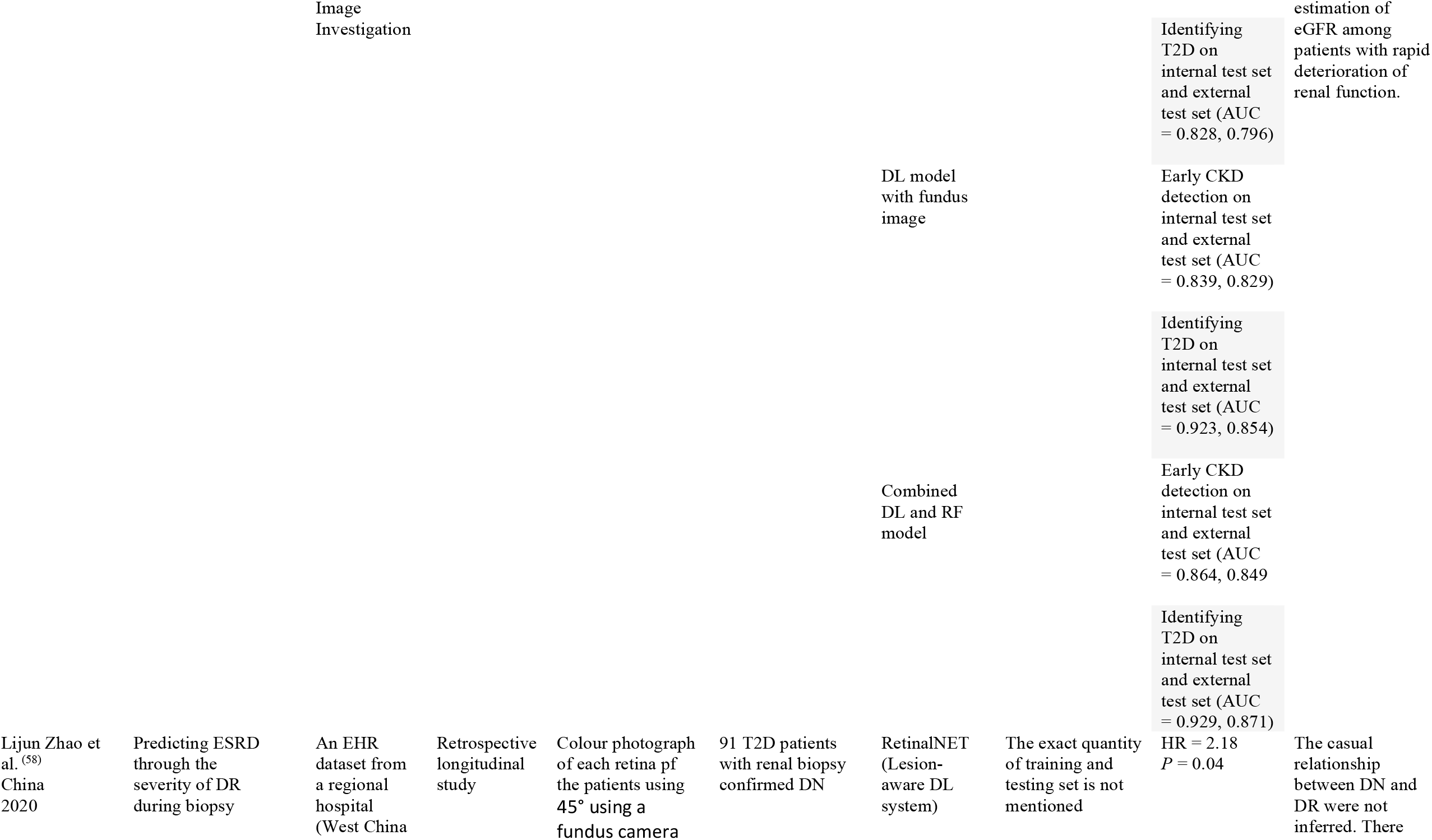
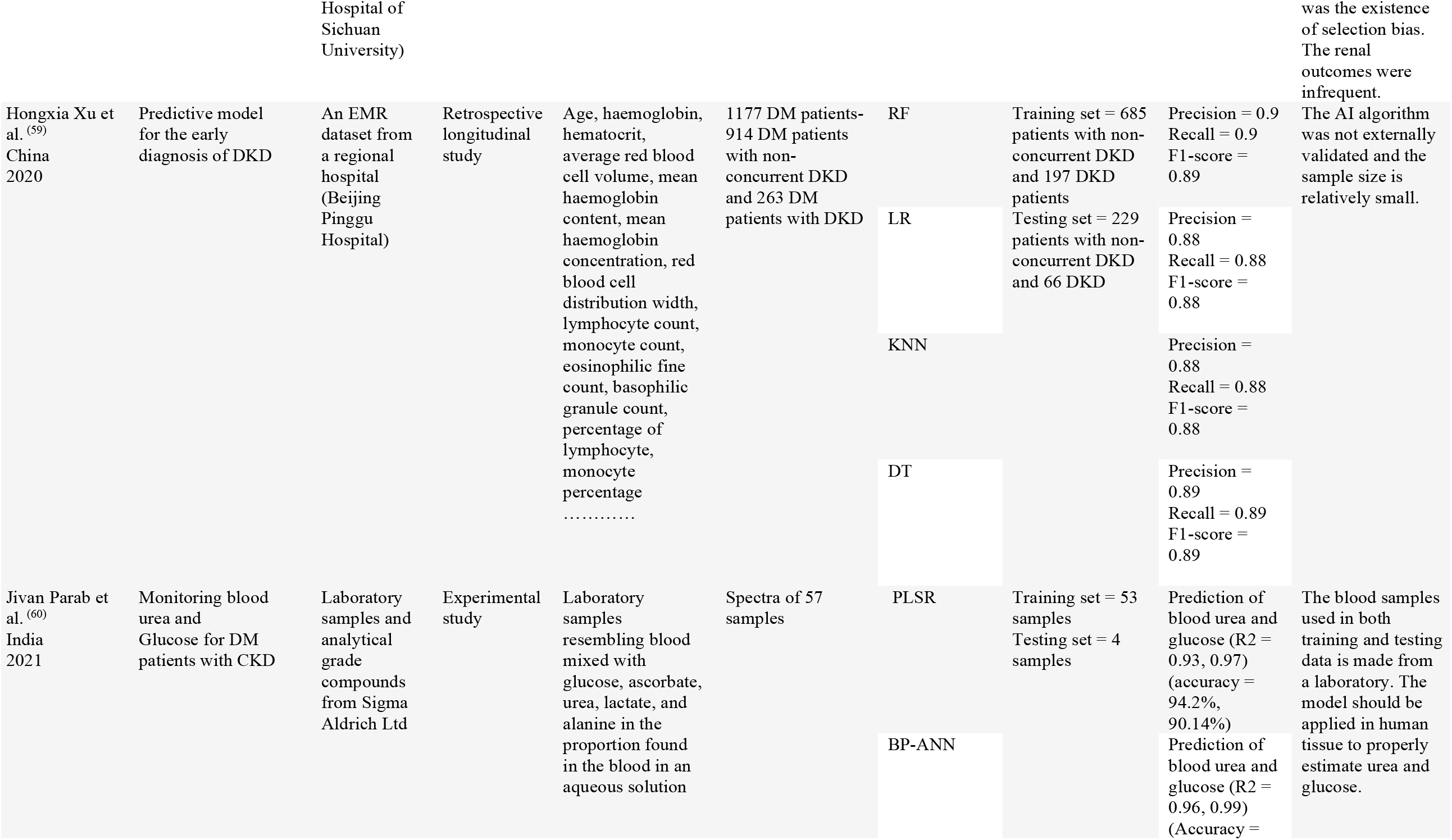
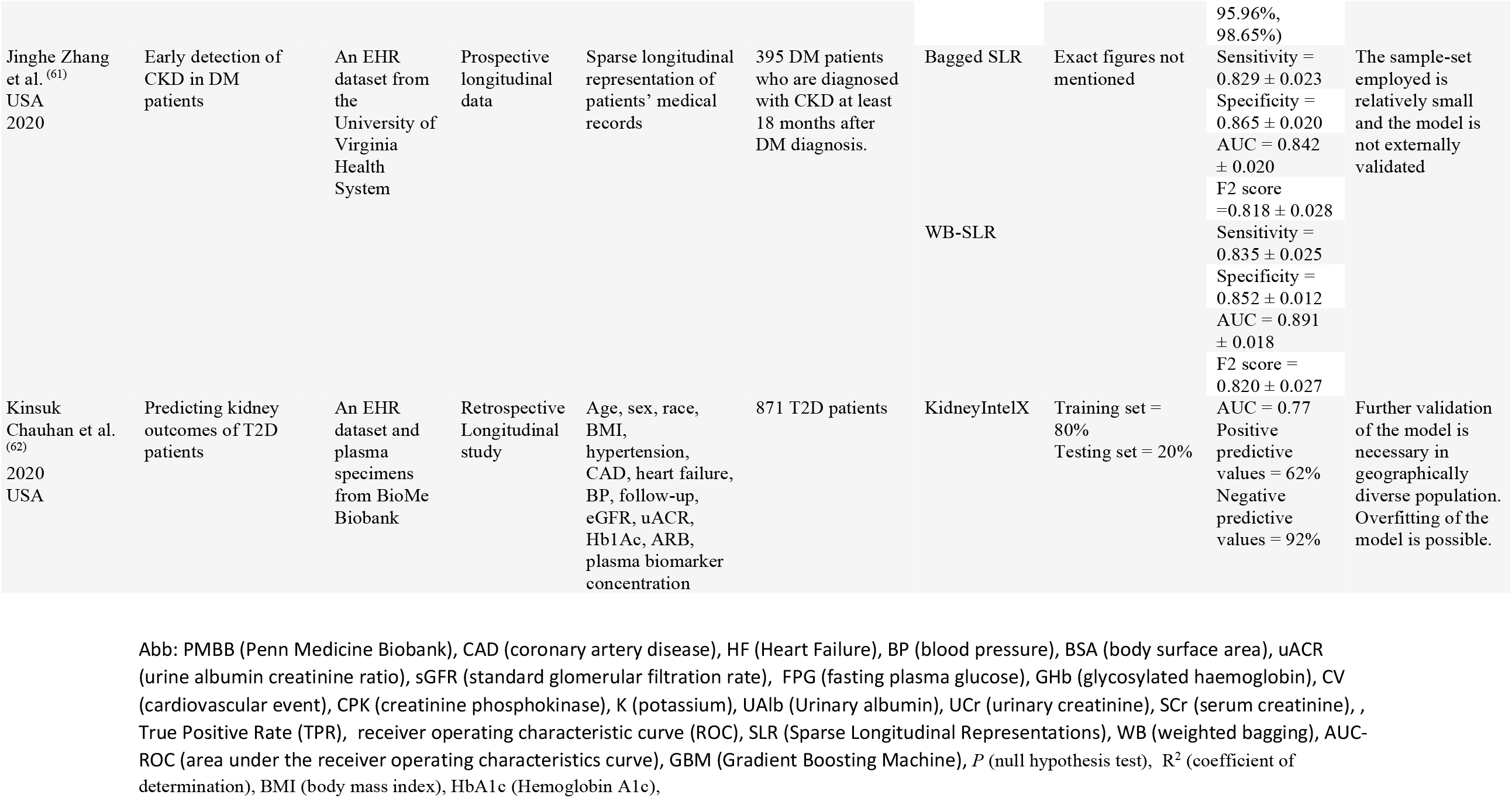
The summary of findings in the included articles

The scheme of this study is summarized into four different categories based on the AI framework in use. The categories include 1) predictive AI model for the early detection of DN (40, 44, 45, 46, 48, 51, 54, 55, 59, 61), 2) AI Framework for the diagnosis of DN (42, 50, 57), 3) predictive model for the progression of DKD (39, 41, 49, 58, 62), 4) the management of DN for existing patients OR early management indication to DM patients without DKD with the help of AI (43, 47, 52, 53, 56, 60). The studies included in this systematic review are from various parts of the world. Seven of the included studies are from the USA (29%), six of them are from China (25%). In addition, three studies are from Japan (12.5%) and Korea (12.5%). Although the number of observational studies included was greater, the comprehensive review consisted of both observational study and experimental study.

A total of 8 studies (39, 40, 44, 45, 48, 49, 53, 59) included a cohort population/ sample size of 1000+ patients/samples to develop the AI model. Meanwhile, 7 studies (41, 50, 51, 52, 54, 55, 57) included a cohort size/ sample size over 10,000 patients/sample size. In addition, a large sum of 14 studies (39, 40, 42, 44, 48, 49, 51, 52, 53, 55, 56, 57, 59, 62) used ML to build the predictive model. On the other hand, 5 studies (47, 50, 54, 58, 60) used DL algorithms as the AI method in their study. Finally, 5 other studies used both ML and DL as separate baselines or in an integrated system (41, 43, 45, 46, 61). It is worth noting that a large number of the studies used EHR datasets from a regional hospital/clinic. A total of nine studies (42, 43, 46, 51, 52, 54, 58, 59, 61) used an EHR dataset from a regional hospital.

Different AI algorithms such as RF (Random Forest), LR (Logistic Regression), SVM (Support Vector Machine), feed-forward network, ANN (Artificial Neural Network), KNN (k-nearest neighbours),), MLP (Multilayer Perceptron), RNN (Recurrent Neural Network), Naive Bayes (NB), Adaboost, DT (Decision Tree), Ridge LR, Apriori, DNN (Deep Neural Network), GBM (Gradient Boosting Machine), convolutional autoencoder, One rule, Simple Logistic, Sequential Minimal Optimizer, LeNet, DetectionNet, Lesion aware DL system, PLSR (Partial Least Square Regression), Backpropagation Artificial Neural Network (BP-ANN). Above all, RF is the most used model for predictive models being used 7 times.

As most of the included studies are predictive models, they require a certain criterion to be met. The data should be pre-processed with the necessary imputation, dimensionality reduction, transformation etc. The training model used in the dataset is used to find the patterns from the feature vectors with the output vectors with the help of the AI algorithm(s). A testing dataset is also necessary from the same source as the training that is unseen to the algorithm. Therefore, the accuracy of the AI algorithm can be assessed. However, a lot of the studies did not mention the exact quantity of the development process of the dataset i.e the training and testing sample size (40 45, 46, 49, 50, 53, 54, 56, 58, 61). As a result, if the performance of the AI model is overfitting or not cannot be determined.

In the quality assessment (Table 4) the shortcomings OR strengths of the studies were assessed. To ensure the included study adds something new to clinical practices and fulfils a void that cannot be assessed by conventional statistical methods, the “limits in current non-AI approach” column is present in Table-4. If the AI model even outperformed the conventional statistical method the cell is filled with a ‘yes’. Except for two studies (59, 45), all 22 other included studies either outperformed conventional methods or accomplished the result which is not attainable through non-AI frameworks. This proves that AI algorithms are superior to conventional methods, and they might be suitable in clinical practices to aid physicians. The feature generation of the AI algorithm(s) before undergoing training is also mentioned. Feature engineering is particularly important as they are essential to select and transform raw data from clinical variables to essential features. As well as, handling missing data or imbalanced data is accounted for in the studies. From the included studies, a total of studies 19 conducted feature engineering in the raw data to make the algorithm more feasible. Four studies (39, 49, 58, 60) did not use feature engineering. Also, before the training of AI algorithms, some studies chose to tune hyperparameters to yield better outcomes. Among 24 studies only four (50, 54, 55, 58) studies did not include hyperparameters in the study. Hyperparameter tuning is necessary for ML models as the optimal combination reduces the predefined lost function as low as possible, which leads to a better performance of the model.

**Table 4:** Quality Assessment of the studies:

|  | Limits in current non-AI approach | Reproducibility |  | External data validation |
| --- | --- | --- | --- | --- |
|  |  | Feature Engineering | Hyperparameters |  |
| Lili Chan et al. <sup>(39)</sup> | Yes | No | Yes | No |
| Hee-Sung Ahn <sup>(40)</sup> | Yes | Yes | Yes | Yes |
| Sunil Belur Nagaraja <sup>(41)</sup> | Yes | Yes | Yes | No |
| Jinxia Chen <sup>(43)</sup> | Yes | Yes | Yes | Yes |
| Xiaoyuan Bao <sup>(44)</sup> | Yes | N/A | Yes | No |
| Baek Hwan Cho <sup>(46)</sup> | Yes | Yes | Yes | No |
| Shinji Kitamura <sup>(42)</sup> | Yes | Yes | Yes | Yes |
| Brandon Ginley <sup>(47)</sup> | Yes | Yes | Yes | Yes |
| Jialing Huang <sup>(48)</sup> | Yes | Yes | Yes | No |
| Lijun Zhao <sup>(58)</sup> | Yes | No | No | Yes |
| Sitong Lin <sup>(56)</sup> | N/A | Yes | Yes | N/A |
| Jivan Parab <sup>(60)</sup> | Yes | No | N/A | No |
| Hongxia Xu <sup>(59)</sup> | No | Yes | Yes | No |
| Kang Zhang <sup>(57)</sup> | Yes | Yes | Yes | Yes |
| Jinghe Zhang <sup>(61)</sup> | Yes | Yes | Yes | No |
| Violeta Rodriguez-Romero <sup>(55)</sup> | Yes | Yes | No | No |
| Xing Song <sup>(51)</sup> | Yes | Yes | Yes | No |
| Xing Song <sup>(52)</sup> | Yes | Yes | Yes | Yes |
| Yoichi Hayashi <sup>(45)</sup> | No | Yes | Yes | No |
| Masaki Makino <sup>(54)</sup> | Yes | Yes | No | No |
| Ji Eun Kim <sup>(50)</sup> | Yes | Yes | No | No |
| Andreas Heinzel <sup>(53)</sup> | Yes | Yes | No | Yes |
| Yong-Mi Kim <sup>(49)</sup> | Yes | No | Yes | No |
| Kinsuk Chauhan <sup>(62)</sup> | Yes | Yes | Yes | No |

The model’s higher predictive accuracy makes it more reliable for clinical use. In addition to the need for high accuracy in the model, validation of the model in different settings outside of the research’s framework is critical. The vast majority of models have not been externally validated. External validation is required to determine the model’s reproducibility and generalizability to unique patients. Aside from that, it is critical to ensure that the model is tested on a multiethnic cohort before implementing the AI framework globally. Only seven (40, 43, 47, 52, 53, 57, 58) of the 24 included articles had the predictive AI model externally validated (29 percent).

## 4. Discussions

This is the first systematic review that reviews current applications of AI in DKD. The heterogeneity in the types of AI algorithms used are shown in the results - SVM, DT, RF, ANN, Naive Bayes etc. The myriads of different feature vectors used in the predictive performances were observed. Studies in this review were performed in different parts of the globe. Some of the studies included multiethnic cohorts. In most of these studies, it is observed that there persist limitations in conventional statistical models. However, ML and DL models proved to be far superior in many instances. AI methods applied in different aspects of DN had met the substantial needs, which conventional methods lack, in the proposed clinical applications. The only aspect of shortcoming in AI algorithms is the generalizability, which can be met with the growing number of EHR datasets in clinics.

It should be pointed out that most selected papers are published within the last four years, which indicates the application of AI in DN is an emerging technology. Despite being not so perfect, AI still prevails a large opportunity to improve the current uses. The predictive models can be improved a lot if expert knowledge is integrated with the data-driven approach. The physicians with adequate knowledge in DN can craft safety constraints as a guide during the training process (16). The performance of AI frameworks in clinical practices of DKD can adequately be assessed from prospective studies (63). For future work, prospective studies should be conducted in studies focused on category four. Therefore, the study would be able to both assess the performance of the AI algorithm along the acceptance and satisfaction of patients with automated approaches.

For the generalizability of the study, the best performing AI algorithm in internal validation should be tested on multiple different ethnic cohorts before adopting its use in clinical practices. On the other hand, further AI devices can be developed that can help the patients of DN to maintain the risk of ESRD at a low rate. However, caution must be adopted before adopting AI algorithms. Such as, whether AI-enabled care is safe and effective enough from the results of robust studies. As well as, the machine-generated results need to be confirmed by a physician or not etc (38).

The review has a few limitations. At the outset, the review only searched open-access databases. Thus, ignoring registers and webpages and other databases such as Scopus and Embase etc. However, this study aimed to only include articles published in peer-reviewed journals to avoid any existing biases. On the other hand, to ensure the reproducibility of the review the manuscript only included open-access databases.

In this study, a systematic effort was made by the author to identify every possible peer-reviewed article available and review AI application in DN. DN is one of the most dangerous complications caused by DM. However, with the proper caution and early detection of DN, a lot of the harmful outcomes can be avoided. The advent of computational biology, with the vast amount of EHR datasets generated every day, enables the further in-depth application of AI in the diagnosis and treatment of DN. Despite its recent emergence in healthcare, they have already outperformed conventional statistical methods. It is high time that physicians adopt AI-based algorithms in clinical approaches of DN.

## Data Availability

All data produced in the present work are contained in the manuscript

